# Public opinions from Malawian and Malawi refugee camp residents of wastewater and environmental surveillance

**DOI:** 10.1101/2024.11.12.24317144

**Authors:** Dammy Jeboda, Brandie Banner Shackelford, Petros Chigwechokha, Brighton A. Chunga, Ayse Ercumen, Cassandra Workman, Joy L. Hart, Ted Smith, Rochelle H. Holm

## Abstract

Across low- and middle-income countries, there have been calls to expand wastewater and environmental surveillance to include non-sewered sanitation systems. Considering public opinion, understanding, and acceptance, as well as any related privacy and personal health information concerns, in this context is important. This study used an in-person survey to learn more about Malawian and Malawi refugee camp residents’ perceptions of wastewater and environmental surveillance as public health tools, and their perceptions of privacy and personal health information. A 15-question survey was conducted from May to July 2024 at three locations in northern, central, and southern Malawi, including a refugee camp (*n* = 536). Some respondents (*n* = 30) also completed a board game and a post-board game survey. The results indicated high public support for surveilling communicable diseases, deadly diseases, environmental toxicants, healthy eating, illegal drugs, mental illnesses, and prescription drugs. Respondents were less supportive of surveillance that may expose their lifestyle behaviors and alcohol use. Regarding sampling locations, the surveillance of an entire city or of camp and schools had the highest acceptance. Some opposition to surveillance in business and religious organizations was found. If their sanitation waste was to be monitored, the respondents wanted the results of the data analysis to be communicated. Our findings suggested that Malawian and Malawi camp residents supported, with little concern, ongoing wastewater and environmental surveillance for public health. Considering privacy thresholds and participant autonomy regarding public health surveillance tools among cultural relevancies is important for future policy development and investment.

## Background

Wastewater and environment-based epidemiology links pathogens and chemicals found in wastewater, the environment, or non-sewered sanitation systems to population-level health.^1^ Wastewater and environmental surveillance (WES) approaches have been used as global public health tools to monitor pathogens, drugs, and dietary patterns. ^2–6^ By using a systematic approach that includes the anonymous and passive sampling of community-generated fecal matter,^1,3,4^ consent is not formally required from the public for wastewater surveillance in United States communities, as the Common Rule definition for human-subject research is not met. While most households in the United States have sewered sanitation systems, in low- and middle-income counties (LMICs), pit latrines and septic tanks are more common^7^ and can also be sampled. Refugee camps in LMICs are particularly vulnerable in terms of having compromised water, sanitation, and hygiene conditions that may facilitate the spread of diarrheal diseases.^8–11^ In LMICs, there has been a call to expand WES.^12^ Since public health interventions, such as vaccination clinics and stay-at-home orders or mandates, informed by WES data would directly impact the community members in LMICs, considering public opinion, understanding, and acceptance, as well as privacy and personal health information (PHI) concerns related to WES, is important.

Within and among cultures, variations exist in how much individuals care about privacy and what information they consider private.^13^ Privacy concerns and expectations are more complex in LMICs lacking centralized waste collection sites and where WES samples may be taken from household pit latrines and septic tanks that have few users. Thus, fecal waste may be considered being closer to stool samples than to pooled samples from a wastewater treatment plant, in high-income countries. Previous research in the United States has generally shown public favorability towards WES,^14,15^ whereas in countries such as Malawi, Africa, there is evidence of strong public opinions surrounding sanitation programs, as well as large-scale resistance and resentment when a community does not feel included in decisions.^16^

WES partially alleviates PHI concerns because specific individuals remain anonymous, as sampling always involves contributions from multiple individuals. However, although WES data do not come directly from an identifiable individual through testing at a clinic or other traditional health settings, this information could still be considered PHI. PHI includes identifiable data related to an individual’s past, present, or future health status.^17^ This point is particularly important, as in many countries across sub-Saharan Africa, culturally, there is less emphasis on an individual and more emphasis on interdependence and community.^18,19^

The notion of WES data as PHI raises important questions concerning how individuals seek to understand the community’s health status. In the context of WES, the collected data may be used to inform and implement policies and other interventions in ways that individuals cannot control. Regarding health research and PHI, concerns can exist regarding how the collected data are used, with whom they are shared, and the desire for control over how the data are released.^20^ Ultimately, however, the primary use of WES data should improve public health.

PHI is an aspect of surveillance that could be important to residents; however, clear PHI regulations may not occur in areas of WES. When individuals worry about PHI being made available to people they do not intend it to, it is a privacy issue and hinges on ethics. Health literacy initiatives by governments and international non-governmental organizations are not always complete.^21^ As in the implementation of community-wide practices and/or policy, researchers are obligated to seek the opinions of stakeholders and to act accordingly. Issues concerning perceptions of PHI as it relates to WES pose potential public concerns for research, data collection, and the analysis of fecal waste-derived data used for guiding public health interventions. Confidentiality in WES may remain a concern for participants if they perceive a risk of re-identification for themselves or their family. Anonymization is crucial in the willingness to share PHI because it can serve to minimize negative outcomes for individuals.^22^ Furthermore, health information is particularly sensitive in a refugee context. A negative human immunodeficiency virus test result was formerly a precursor to resettlement in the United States.^23^ In the United States, discussions surrounding wastewater surveillance under the Fourth Amendment concluded that there is no reasonable expectation of privacy in instances of wastewater collection and analysis from public sewers.^24^

To date, only four WES public opinion surveys have been developed, with each conducted in the United States focusing on sewered sanitation systems.^14,15,25,25^ The objective of this survey was to utilize survey data to identify public opinion and privacy concerns, in order to better understand how WES is accepted as a public health tool in an LMIC setting where non-sewered sanitation systems are dominate. The results can help inform WES scale-up strategies for wastewater and environmental surveillance in both urban and camp locations to improve community health.

## Methods

### Study sites

For comparative purposes, our study included three sites: Mzuzu City in northern Malawi; Goliati in Traditional Authority (TA) Chimaliro in southern Malawi; and Dzaleka Refugee Camp in Dowa District, central Malawi (Figure 1). These sites were chosen to represent different settings in Malawi. Mzuzu has a population of 221,272.^27^ It has a large regional referral hospital, serves as a commercial hub for the region, and hosts Mzuzu University, a government institution of higher education. There were two survey sites in Mzuzu: 1.) Mzuzu University, and 2.) the community area. TA Chimaliro is located in Thyolo District and has a population of 56,764.^27^ It is characterized as being mostly a rural farming area, is located 1 h from most commerce and healthcare facilities, and hosts Malawi University of Science and Technology, a government institution of higher education. Dzaleka Refugee Camp is located in the Dowa district and is the only camp for refugees and asylum seekers in Malawi. As of July 2024, the camp was hosting over 54,000 refugees and asylum seekers, most of whom came from the Democratic Republic of Congo (65%), Burundi (22%), and Rwanda (13%). The camp is overcrowded, food-insecure, and water-scarce and has an under-resourced health system. Dzaleka Refugee Camp represents a unique opportunity for WES public-opinion research because the sample collection for WES is highly visible to residents and all non-sewered sampling; before sampling, community sensitization was conducted to increase the awareness of those living in the camp of the WES effort. Uniquely, each of the three locations studied had active WES sampling during the survey.

**Figure 1:**
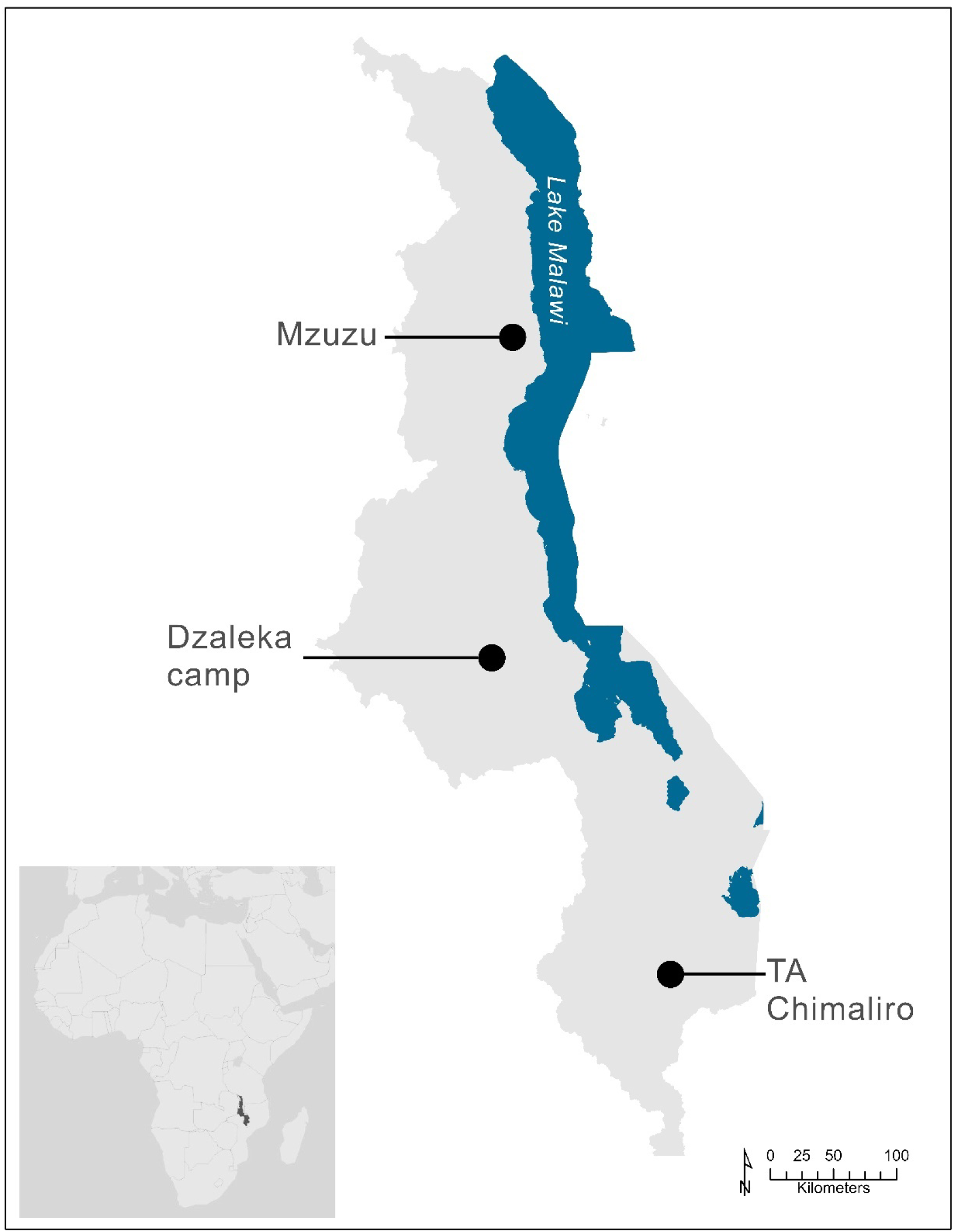
Study sites

### Sample recruitment

Data for the analysis (n = 536) were collected in-person from May to July 2024. Considering that the nationwide population is approximately 20 million, a minimum sample size of 500 was selected to ensure stable correlations with 95% confidence.^28^ The enrollment targets were 350 respondents from Mzuzu, 100 from TA Chimaliro, and 50 from Dzaleka. Actual enrollment at the three study sites differed from the targeted enrollment, based on logistical constraints; however, a sample size of >500 was maintained for the study. Adult respondents were recruited from the study sites via convenience sampling, they were approached by a researcher to complete the surveys in English or Chichewa. At Dzaleka, survey recruitment was limited to an area near the entrance, designated for people who had not yet received a shelter plot. Survey respondents were limited to adults of 18 years old or older. A convenience subset of respondents (*n* = 30), based on time available, from either Mzuzu or TA Chimaliro also completed a board game and a post-board game survey.

### Survey

The goal of the 15-question survey was to learn more about the public’s perceptions of WES as a public health tool and surrounding privacy issues. The survey questions were adapted from a WES survey previously used in the United States,^15^ designed to assess multiple aspects of public opinion, such as comfort with WES, accepted surveillance target parameters and geographic scale, and privacy concerns. The respondents also answered questions regarding their age, gender, and education. Additional items were added to assess the toilet type used most often, perceptions of health data confidentiality, and importance of autonomy over PHI data. Surveys were conducted on paper with the aid of Chichewa translators, as needed. The survey questions were either read aloud by the translator or completed by the participants. The survey completion time ranged between 10 and 20 min. The full survey instrument is shown in Supplemental Material Text S1.

When asked to indicate their level of comfort with the nine WES biological and chemical surveillance targets, participants did so on a Likert scale ranging from 1 (very uncomfortable) to 5 (very comfortable). The respondents were asked to indicate any geographic scale(s) at which they wanted sanitation system waste to be monitored. Response options included the entire city/camp monitored in a more anonymous manner, and/or neighborhoods, businesses, prisons, schools, or houses. The respondents were also asked if any areas or geographic scale(s) should be prohibited from surveillance, and regarding sanitation system waste monitoring as an assessment of invasion of privacy. To assess trust, respondents were asked to what extent they would support various groups accessing their health data, on a Likert scale ranging from 1 (full opposition) to 5 (full support). Respondents were also asked to rate from 1 (unconcerned) to 5 (very important) the importance of their health data results being communicated back to them and of the data remaining anonymous/protected. When asked to indicate their confidence in officials in keeping personal information private, and how much of their privacy they were willing to give up for others, participants also answered on a Likert scale ranging from 0 (no confidence) to 4 (complete confidence). Respondents also ranked five health topics as the most important to them (1, ‘most important’; 5, ‘least important’). To capture the concerns or sentiments not addressed in the survey, respondents were asked a final open-ended question regarding whether they had any concerns or comments about WES samples being collected from pit latrines and septic tanks in the area. No incentives were provided to participants.

### Board game

Contingent valuation is an economic approach used in surveys that creates a hypothetical market and asks respondents to consider their willingness to pay (how much a respondent is willing to pay to acquire a non-market good) and/or willingness to accept (the amount of compensation the respondent is required to give up), based on the existence of this market.^29,30^ This study employed contingent valuation, creating hypothetical scenarios to investigate the willingness of participants to give up their privacy in exchange for personal and community health gains. Generally, individuals choose to relinquish some of their rights and provide personal information in exchange for services that may improve their health. Conversely, in this board game, by retaining privacy, individuals may miss potential benefits. Acquisti et al.^13^ highlighted the importance of context when individuals decide whether to retain their privacy.

In this study, the board game (Figure 2) aimed to help respondents engage creatively with WES principles. Players considered scenarios that tested their willingness to give up their PHI for personal and community health incentives, while weighing the negative outcomes. After receiving the instructions, the participants played the board game individually. Players were assigned privacy (100%), personal health (30%), and community health (40%) in the form of game tokens. In each of seven rounds (Table 1), participants were presented with an offer from a private research group that they were told would outsource the data they provided in order to offer incentives. The incentives would increase their personal and/or community health information in varying amounts in exchange for 10% of their privacy in each round. Participants were encouraged to act as they would if the scenarios were real and to decide to either accept the offer proposed by the private research group and gain incentives or reject the offer and end the board game. After being given the opportunity to ask clarifying questions, the researcher (JD) explained the scenario for each round. The researcher recorded the final board game outcomes for each participant. The post-board game survey included three open-ended questions used to further investigate the decision-making process of participants during the board game.

**Figure 2.**
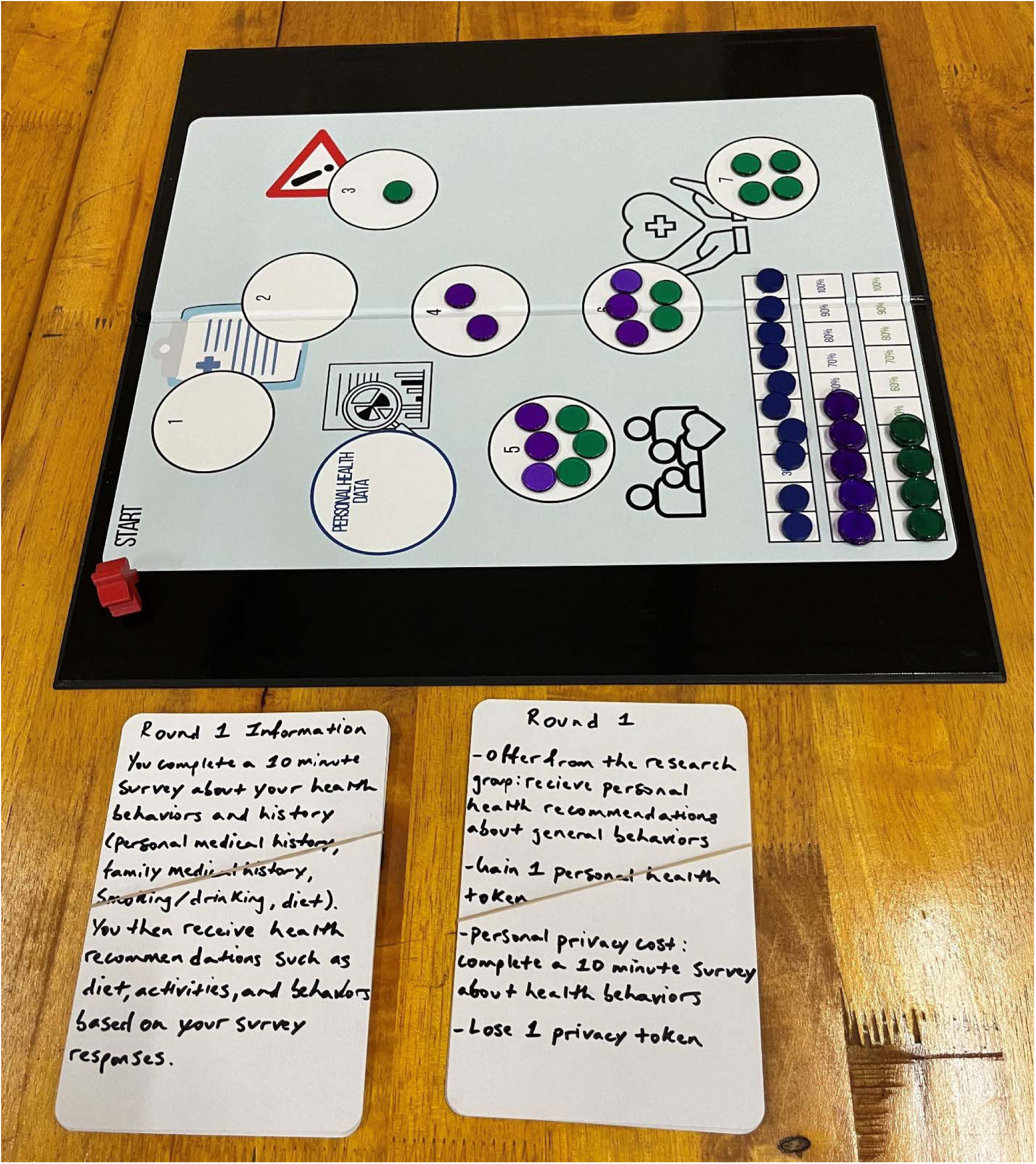
Board-game-based contingent valuation used in this study to investigate public opinion about wastewater and environmental surveillance.

**Table 1.** Board-game-based contingent valuation used in this study to investigate privacy and willingness to give up personal health information for personal and community health incentives over seven rounds.

| **Privacy given up** | **Personal health** | **Community health** |
| --- | --- | --- |
| Round 1<br>Offer: Participants complete a 10-min survey about health behaviors<br>- (personal medical history, family medical history, medication history and adherence, smoking/drinking, and diet)<br>-Cost: 1 privacy token | Round 1<br>-Receive personal health recommendations about general behaviors<br>-Gain: 1 personal health token | Round 1<br>-None |
| Round 2<br>-Wastewater is sampled at the city level and tested<br>-Cost: 1 privacy token | Round 2<br>-Receive alerts about disease outbreaks detected in wastewater<br>-Receive personal health recommendations (health status, disease risk, etc.)<br>-Gain: 3 personal health tokens | Round 2<br>-None |
| Round 3<br>-Researchers have a narrower focus for wastewater monitoring and know which data came from their community<br>-Cost: 2 privacy tokens | Round 3<br>-None | Round 3<br>-Wastewater monitoring occurs at the community level<br>-Gain: 1 community health token |
| Round 4<br>-Complete another, more in-depth survey and lab tests such as blood pressure, heart rate, and body mass index measurements<br>-Cost: 1 privacy token | Round 4<br>- Receive in-depth personal health recommendations (health status, and risk of disease)<br>-Gain: 2 personal health tokens | Round 4<br>-None |
| Round 5<br>-Data is shared with additional groups<br>-Cost: 1 privacy token | Round 5<br>-Data previously given is shared with (non-specified) outside groups to gain info about personal and community health and make a wider variety of recommendations<br>-Gain: 3 personal health tokens | Round 5<br>-Data previously given is shared with (non-specified) outside groups to gain info about personal and community health and make a wider variety of recommendations |
|  |  | -Gain: 3 community health tokens |
| Round 6<br>-Researchers know which household the wastewater they are monitoring comes from<br>-Cost: 1 privacy token | Round 6<br>-Wastewater samples are collected at the household level<br>-Gain: 3 personal health tokens | Round 6<br>-Wastewater samples are collected at the household level<br>-Gain: 2 community health tokens |
| Round 7<br>-Data are published in a database that other groups (e.g., government, and private research groups) can access/use without additional consent from the participant<br>-Cost: 1 privacy token | Round 7<br>-None | Round 7<br>- Potential positive consequences from making their data available<br>-Gain: 4 community health tokens |

### Analysis

The respondents from which the survey data were physically collected by the researcher (JD) at the camp were classified as “Dzaleka.” The respondents who reported using a pit latrine as their most common household toileting facility were classified as “pit latrine users.”

The survey results were entered manually into Excel. Quantitative data analyses for the board game actions and survey answers were performed using R version 4.3.1 and R Studio 2023.06.1+524.^31^ The analysis focused on describing the differences in the public opinion of Malawian or Malawi camp residents regarding WES within 95% confidence intervals (CIs). A data-driven approach was used to identify qualitative themes by distilling the qualitative survey data, identifying/comparing themes, and creating codes.^32^ The responses were coded into ten qualitative themes: no concern; spread/contraction of disease or pollution; concern for sample technician safety; extractive research practices; awkward/inappropriate/uncomfortable research practices; contributions to community health; contributions to personal health; skepticism or mistrust of researchers/research practices, or mishandling of PHI; concern with the current state of the toilet facility; and consent.

### Ethics approval and consent to participate

This study was reviewed and approved by the Malawi University of Science and Technology Research Ethics Committee (P.01/2024/117) and the University of Louisville Institutional Review Board (24.0245). Written informed consent was obtained from all participants.

## Results

### Description of Survey Population

This analysis included 536 respondents, with complete data for all questions. Of the 536 respondents, 80% (*n =* 429) were located in Mzuzu, 11% (*n =* 57) in Dzaleka, and 9% (*n =* 50) in TA Chimaliro. More than half of the respondents (306/536; 57%) reported using a pit latrine most often (referred to as “pit latrine users” hereafter) at their home, with the remaining nearly 42% (224/536) using a flush toilet to a septic tank and 1% (6/536) using no facility/open defecation. The respondents were predominantly male (344/536; 64%), aged between 18 and 34 years (415/536, 77%), and without a university or technical degree (409/536; 76%). The demographic characteristics of the respondents were stratified by groups of total survey respondents, Dzaleka camp residents, and pit latrine users and are shown in Table 2.

**Table 2.** Demographic characteristics of respondents.

|  | Total survey<br>*n* (%) | Dzaleka camp resident<br>*n* (%) | Pit latrine user<br>*n* (%) |
| --- | --- | --- | --- |
| **Gender** |  |  |  |
| Female | 190 (35.4%) | 38 (66.7%) | 107 (35%) |
| Male | 344 (64.2%) | 18 (31.6%) | 197 (64.4%) |
| Other | 1 (0.2%) | 1 (1.8%) | 1 (0.3%) |
| Prefer not to say | 1 (0.2%) | 0 (0%) | 1 (0.3%) |
| **Age** |  |  |  |
| 18-24 | 216 (40.3%) | 10 (17.5%) | 118 (38.6%) |
| 25-34 | 199 (37.1%) | 20 (35.1%) | 120 (39.2%) |
| 35-44 | 78 (14.6%) | 12 (21.1%) | 40 (13.1%) |
| 45-54 | 33 (6.2%) | 9 (15.8%) | 20 (6.5%) |
| 55-64 | 7 (1.3%) | 5 (8.8%) | 7 (2.3%) |
| 65-74 | 1 (0.2%) | 1 (1.8%) | 1 (0.3%) |
| 75-84 | 2 (0.4%) | 0 (0%) | 0 (0%) |
| **Highest education level** |  |  |  |
| None | 14 (2.6%) | 14 (24.6%) | 14 (4.6%) |
| Primary School | 56 (10.4%) | 23 (40.4%) | 52 (17%) |
| Secondary School | 159 (29.7%) | 18 (31.6%) | 111 (36.3%) |
| Some University | 180 (33.6%) | 1 (1.8%) | 96 (31.4%) |
| University Degree | 23 (4.3%) | 1 (1.8%) | 23 (7.5%) |
| Technical Degree | 104 (19.4%) | 0 (0%) | 10 (3.3%) |
| **Toilet used most often** |  |  |  |
| Pit latrine | 306 (57.1%) | 57 (100%) | 306 (100%) |
| Flush toilet to septic tank | 244 (45.5%) | 0 (0%) | 0 (0%) |
| No facility/open defecation | 6 (1.1%) | 0 (0%) | 0 (0%) |

### Total survey trends

Respondents were overwhelmingly supportive of the nine WES targets presented in the survey (Figure 3). In the total survey, participants were more likely to support monitoring for healthy eating (mean Likert score = 4.5; 95% CI = 4.4, 4.6), communicable diseases (mean = 4.4; CI = 4.3, 4.5), environmental toxicants (mean = 4.4; CI = 4.3, 4.5), deadly diseases (mean = 4.4; CI = 4.3, 4.5), prescription drugs (mean = 4.4; CI = 4.3, 4.5), illegal drugs (mean = 4.3; CI = 4.1, 4.4), and mental illnesses (mean = 4.2; CI = 4.1, 4.3). Less support was found for lifestyle behaviors (e.g., smoking, and birth control) (mean = 4.0; CI = 3.9, 4.1) and alcohol consumption (mean = 3.9; CI = 3.8, 4.1).

**Figure 3.**
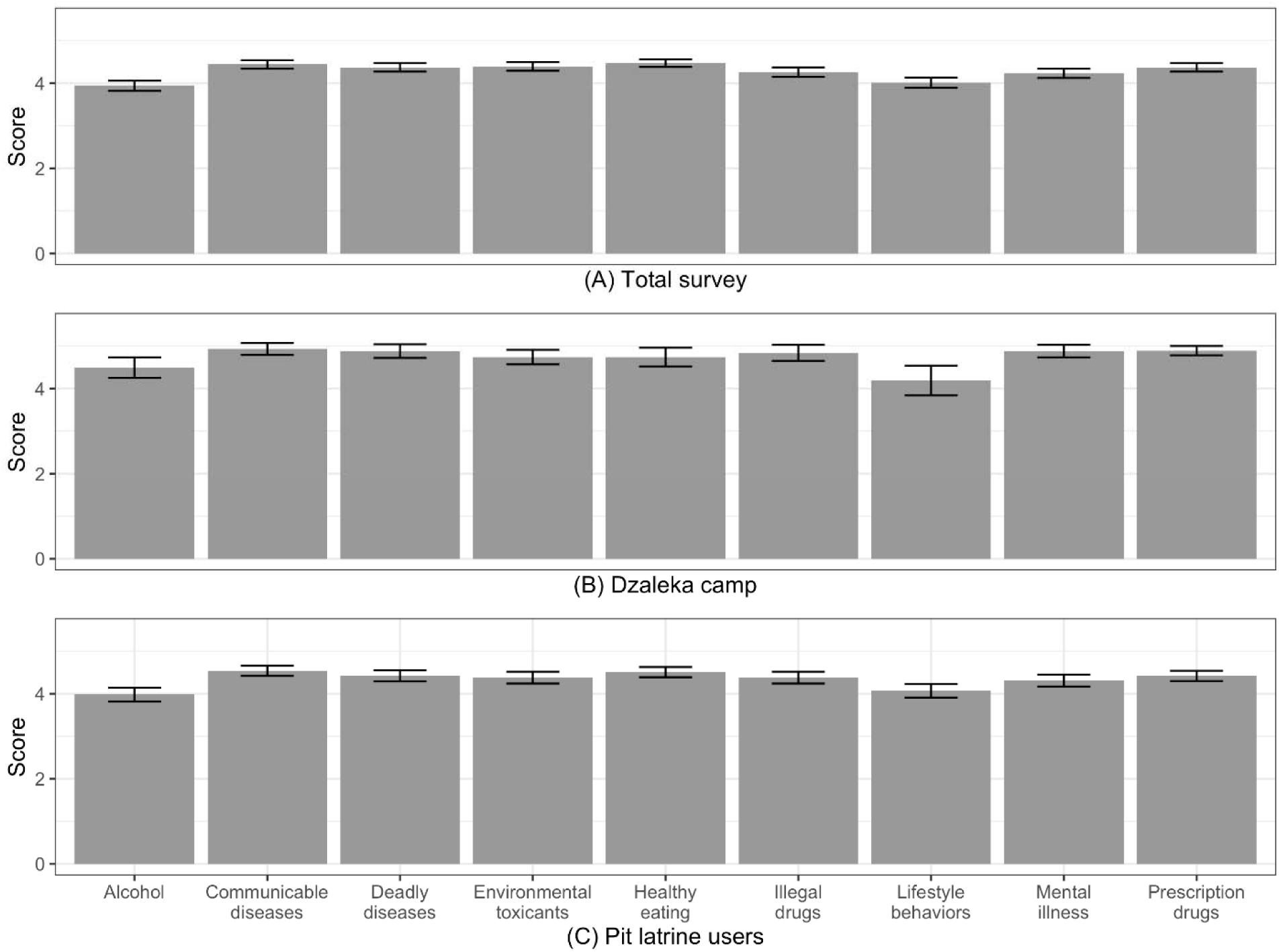
Summary of public opinion on level of comfortability across biological and chemical targets for wastewater and environmental surveillance. Participants ranked on a Likert scale from 1 (very uncomfortable) to 5 (very comfortable) for each of the nine targets. Error bars represent 95% confidence intervals. Panel A) Total survey (*n =* 536); B) Subset of total respondents located in Dzaleka camp (*n =* 57); C) Subset of total respondents that were pit latrine users (*n =* 306).

Respondents were also supportive of the six geographic sampling scales that WES could hypothetically cover in pooled sampling (Figure 4). Near-universal support was provided for school monitoring (95%; *n =* 508). High levels of support were provided for the other geographic categories: monitoring an entire city or camp (87%; *n =* 468), prisons (87%; *n =* 465), houses (86%; *n =* 462), neighborhoods (82%; *n =* 438), and businesses (79%; *n =* 426). More than half (62%) of the total survey respondents supported monitoring each of the six location options. Similarly, when respondents were asked whether sanitation system waste monitoring should be prohibited in any of the four areas, few oppositions were noted. The opposition to monitoring in religious organizations (12%; *n =* 62) was higher than that in private companies (7%; *n =* 39), individual households (5%; *n =* 26), and schools/colleges/universities (2%; *n =* 12). For the two questions, schools demonstrated the highest support for and least opposition to WES.

**Figure 4.**
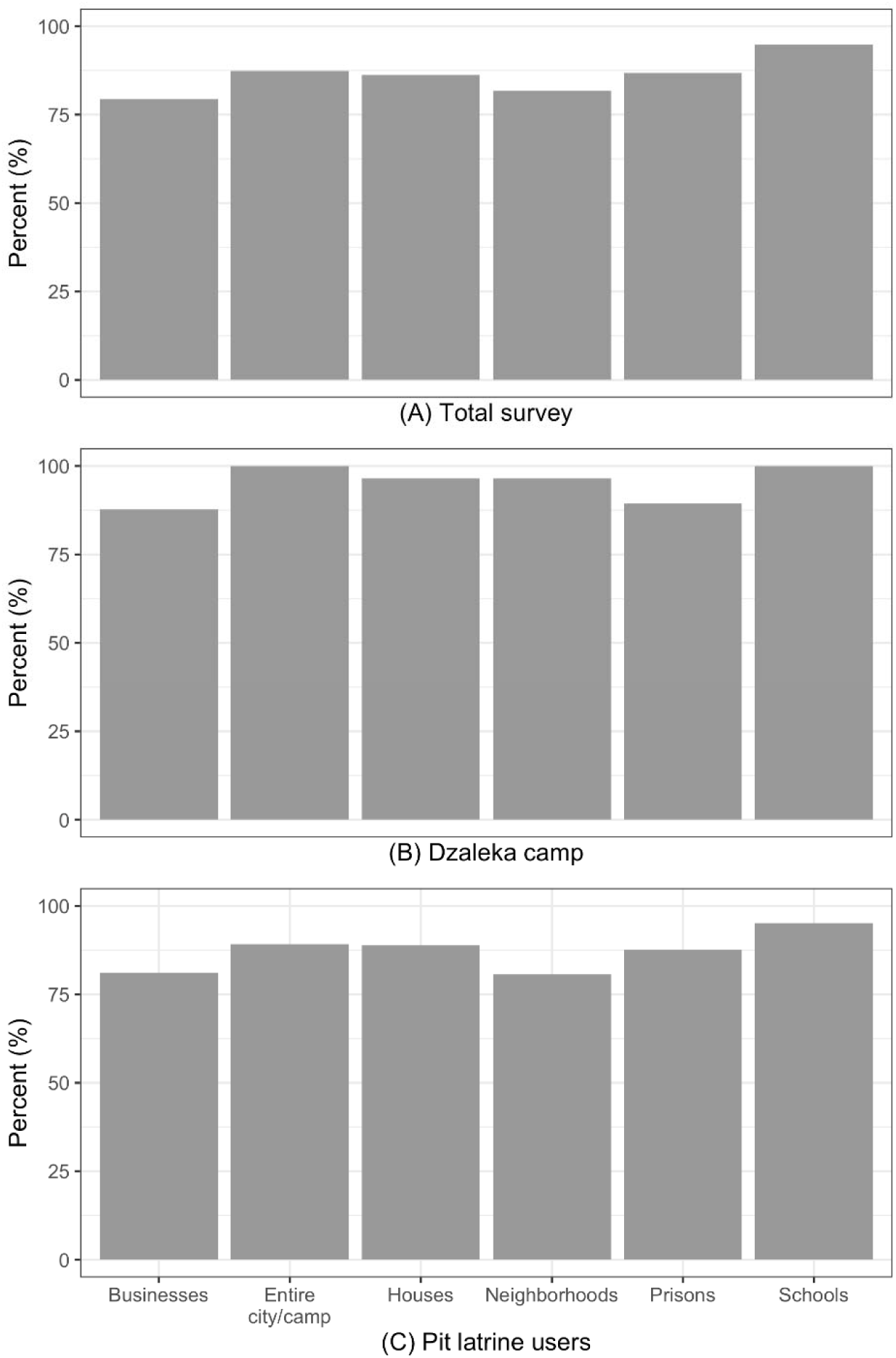
Percentage of public opinion supporting the sampling of wastewater and environmental surveillance at a range of six geographical sampling scales. Respondents were asked to indicate (yes/no) the geographic scale(s) at which they would want sanitation system waste monitored. Panel A) Total survey (*n =* 536); B) Subset of total respondents located at Dzaleka camp (*n =* 57); C) Subset of total respondents that were pit latrine users (*n =* 306).

When asked if conducting sanitation system waste monitoring was an invasion of privacy, the majority (75%; *n =* 401) of the total survey respondents indicated that it was not.

Government officials (mean Likert score = 4.4; CI = 4.3, 4.5) and academic researchers (mean = 4.4; CI = 4.3, 4.5) were the most trusted in accessing health data, followed by consultant research groups (mean = 4.2; CI = 4.2, 4.3) and NGO-supported researchers (mean = 4.2; CI = 4.1, 4.3) (Figure 5). The lowest support was for the block leader (a municipally elected community leader) accessing respondents’ health data (mean = 3.7; CI = 3.6, 3.9). Respondents had the most confidence in officials keeping their health or medical information private (mean = 3.3/4; CI = 3.2, 3.4), and lower confidence in providing lifestyle/behavior information (mean = 2.2/4; CI = 2.1, 2.3) and financial information (mean = 2.1/4; CI = 1.9, 2.2).

**Figure 5.**
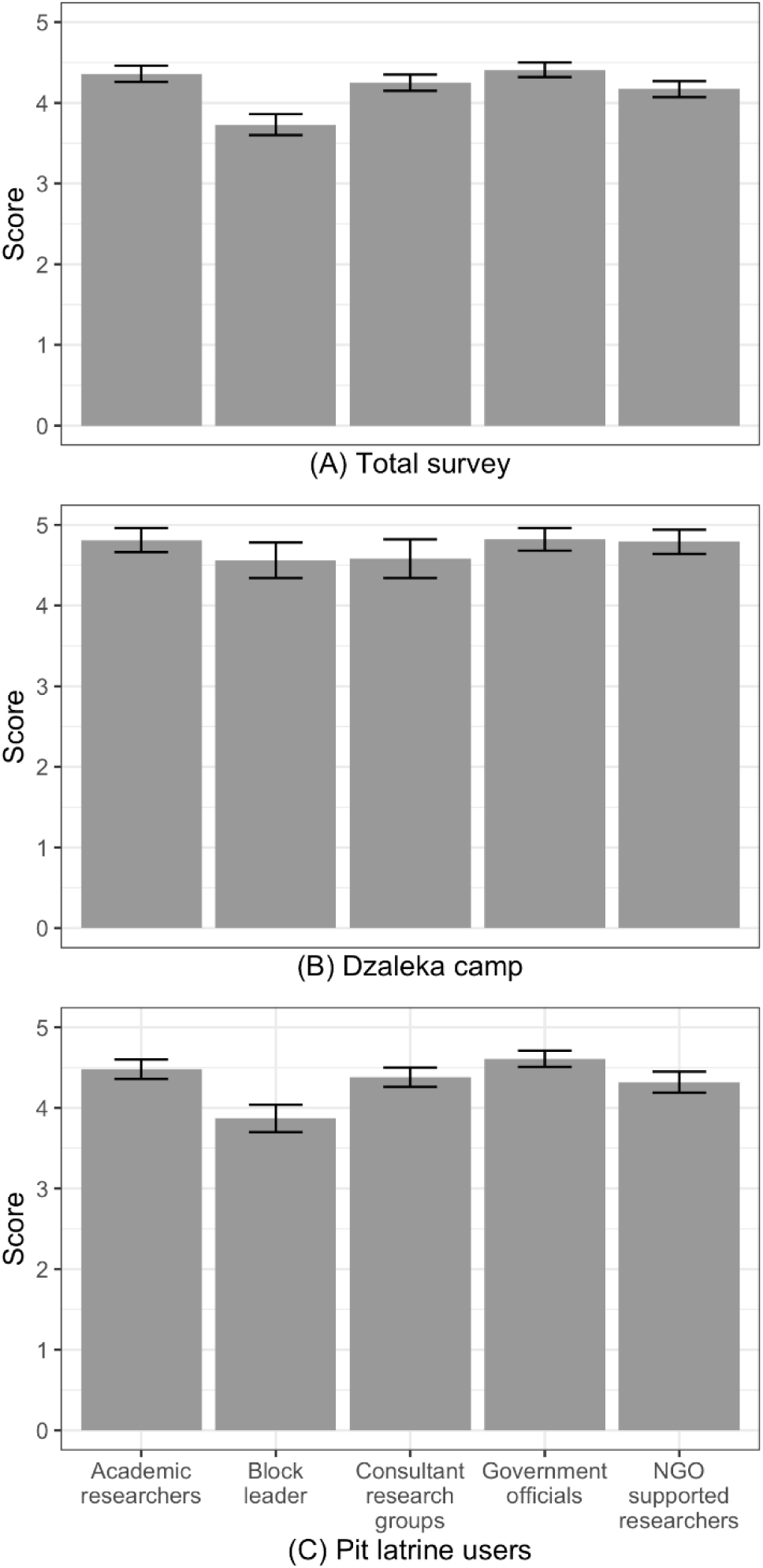
Summary of public opinion for trust regarding accessing health data across five groups of local decision makers. Respondents were asked to indicate confidence in listed groups accessing health data, on a Likert scale ranging from 1 (full opposition) to 5 (full support). Panel A) Total survey (*n =* 536); B) Subset of total respondents located at Dzaleka camp (*n =* 57); C) Subset of total respondents that were pit latrine users (*n =* 306).

When respondents ranked health topics by importance, the most important (1 is ‘most important’ and 5 is ‘least important’) was to protect ‘my’ health (mean = 1.6; CI = 1.5, 1.7), followed by to ‘protect the health of my loved ones’ (mean = 2.2; CI = 2.1, 2.3), ‘protect the health of people I know’ (mean = 3.2; CI = 3.1, 3.3) and ‘protect the health of people in city/camp’ (mean = 3.6; CI = 3.5, 3.7). The least important was ‘protecting the health of people I do not know’ (mean = 4.4; CI = 4.4, 4.5).

When asked how much privacy respondents would be willing to give up to ensure that people in their area could live safe and healthy lives, with 0 indicating none and 4 indicating all, respondents were most willing to give up their health/medical information (mean = 3.0/4; CI = 2.9, 3.1) and lifestyle/behavior information (mean = 2.5/4; CI = 2.4, 2.7). However, respondents were least willing to give up privacy related to their financial information (mean = 1.9/4; CI = 1.8, 2.1).

Most respondents (97%; *n =* 520) indicated that it was important or very important for data to be communicated back. Respondents were also concerned (77% important or very important; *n =* 414) regarding their health data remaining anonymous/protected.

### Dzaleka Refugee Camp trends

The Dzaleka survey site was unique in that nearly all respondents were aware of WES practices in the camp, having recognized the sample collectors’ work attire and personal protective equipment during collection. Compared to the total survey, this subset of camp respondents was more likely to support monitoring across the nine WES targets presented in the survey, but the rank where support was highest differed from that of the total survey. Nearly all the camp resident respondents supported the monitoring of communicable diseases (mean = 4.9; CI = 4.8, 5.1), whereas no target received this high of total survey support. High support was also found for prescription drug categories (mean = 4.9; CI = 4.8, 5), mental illnesses (e.g., monitoring the presence of stress hormones; mean = 4.9; CI = 4.7, 5), deadly diseases (mean = 4.9; CI = 4.7, 5), illegal drugs (mean = 4.8; CI = 4.6, 5), environmental toxicants (mean = 4.7; CI = 4.6, 4.9), and healthy eating (mean = 4.7; CI = 4.5, 5). Similar to the total survey, the lowest two categories of support were for alcohol consumption (mean = 4.5; CI = 4.3, 4.7) and lifestyle behaviors (mean 4.2; CI = 3.8, 4.5); however, support remained higher when compared to the total survey trends for these categories.

Regarding the six geographic scales that WES could cover, all camp residents surveyed supported surveillance for the entire city or camp (100%; *n =* 57) and schools (100%; *n =* 57), while support for neighborhoods (96%; *n =* 55) and houses (96%; *n =* 55) was high. Prisons (89%; *n =* 51) and businesses (88%; *n =* 50) were less supported. When respondents were asked whether sanitation system waste monitoring should be prohibited in any of the four areas, there was no opposition (0%, *n =* 0) for either schools/colleges/universities or individual households. Only one respondent was opposed to surveillance at private companies (2%; *n =* 1), and a few respondents were opposed to surveillance at religious organizations (9%; *n =* 5). Pertinently, the camp respondents did not appear to oppose monitoring for the entire camp, nor if it was conducted across neighborhoods, schools, or households.

When asked if conducting sanitation system waste monitoring was an invasion of privacy, fewer camp respondents than that for the total survey (25% for the total survey versus 18% for camp residents) indicated that it was. Nearly all camp resident respondents (98%; *n =* 56) indicated that it was important or very important that data be communicated back, similar to the total survey results. Compared to the total survey, respondents were less concerned (52% important or very important; *n =* 29) about their health data remaining anonymous/protected.

### Pit latrine user trends

The respondents who indicated that they used pit latrines (*n* = 306) most often may have been from any of the three survey locations. All Dzaleka camp residents were pit latrine users; however, considering camp residents in combination with Mzuzu and TA Chimaliro, respondents allowed for further investigation, as pit latrine pooling of individual stool might have been less if, for example, samples were collected from the top of a pit latrine.^33^ Pit latrine users were again highly supportive of analysis targets including communicable diseases (mean = 4.5; CI = 4.4, 4.7), healthy eating (mean = 4.5; CI = 4.4, 4.6), deadly diseases (mean = 4.4; CI = 4.3, 4.5), prescription drugs (mean = 4.4; CI = 4.3, 4.5), illegal drugs (mean = 4.4; CI = 4.2, 4.5), environmental toxicants (mean = 4.4; CI = 4.2, 4.5), and mental illnesses (mean = 4.3; CI = 4.2, 4.5). Similar to the total survey and the subset of the Dzaleka camp survey respondents, less support was indicated for lifestyle behavior (mean = 4.1; CI = 3.9, 4.2) and alcohol consumption (mean = 4; CI = 3.8, 4.1) surveillance.

Regarding public opinion supporting the sampling of WES at a range of six geographical sampling scales, similar to the total survey and the Dzaleka camp survey, pit latrine users again showed schools having the highest support (95%; *n =* 291), followed by the entire city or camp (89%; *n =* 273), houses (89%; *n =* 272), and prisons (88%; *n =* 268). Lower support was indicated for neighborhoods (81%; *n =* 247) and businesses (81%; *n =* 248). A few respondents opposed monitoring in schools (3%; *n =* 8), households (4%; *n =* 11), and private companies (5%; *n =* 15). Similar to the total survey and the subset of the Dzaleka camp survey results, some respondents opposed surveillance at religious organizations (12%; *n =*38).

When asked whether conducting sanitation system waste monitoring was an invasion of privacy, many pit latrine users (76%; *n =* 233) indicated that it was not. Most respondents (96%; *n =* 295) again indicated that it was important or very important that, if sanitation waste was to be monitored, the data would be communicated back to them. Respondents were more concerned (73% important or very important; *n =* 223) about their health data remaining anonymous/protected compared to the concern at the camp, which was similar to the total survey trends.

### Privacy

For respondents who indicated that they viewed sanitation system waste monitoring as an invasion of privacy (*n =* 87), further analysis was conducted to understand these privacy concerns. The mean was lower although similar among this group in terms of willingness to give up health/medical information privacy to ensure that others in their area could live healthy lives (mean of = 2.88/5 verses 3.0/5 in the total survey), as well as to give up lifestyle/behavior information (mean of 2.2/5 versus 2.5/5 in the total survey); however, this group was slightly more willing to give up their privacy surrounding financial information (mean of 1.97/5 versus in the total survey). The importance of health data remaining anonymous/protected in WES was similarly high in this group and in the total survey, at 75% (*n =* 87) and 77% (*n =* 214), respectively. There was also the most support in this group for government officials accessing health data (mean 4.15/5), and the least support for local block leaders (mean 3.3/5), which was similar to the trust trend for the total survey.

### Comments or concerns about samples being taken

Qualitative analysis of the total survey data indicated that most (80%; *n =* 431) respondents did not have any comments or concerns about WES sampling in their area. Of the six respondents (1%) who reported having no household toilet facility or practicing open defection, each reported no comments or concerns about samples being taken; these respondents would not reasonably be expected to be covered by WES sampling in their households. Of the respondents who had concerns or comments (20%; *n =* 105) (Table 3), approximately 40% (*n =* 42) provided positive feedback about the sample collection contributing to community health, and 3% commented on the sample taking improving their personal health. The most common respondent-related negative concerns (15%; *n =* 16) cited mistrust or skepticism of research practices, researchers themselves, or information remaining private. In addition, a group of concern themes was related to the logistics of sample collection, in reference to the current state of their toilet facility (10%; *n =* 11), the spread/contraction of diseases or pollution from sampling activities (9%; *n =* 9), or the safety of the sample collectors (5%; *n =* 5). Nine percent of the respondents (*n =* 9) were concerned about extractive research practices.

**Table 3.**
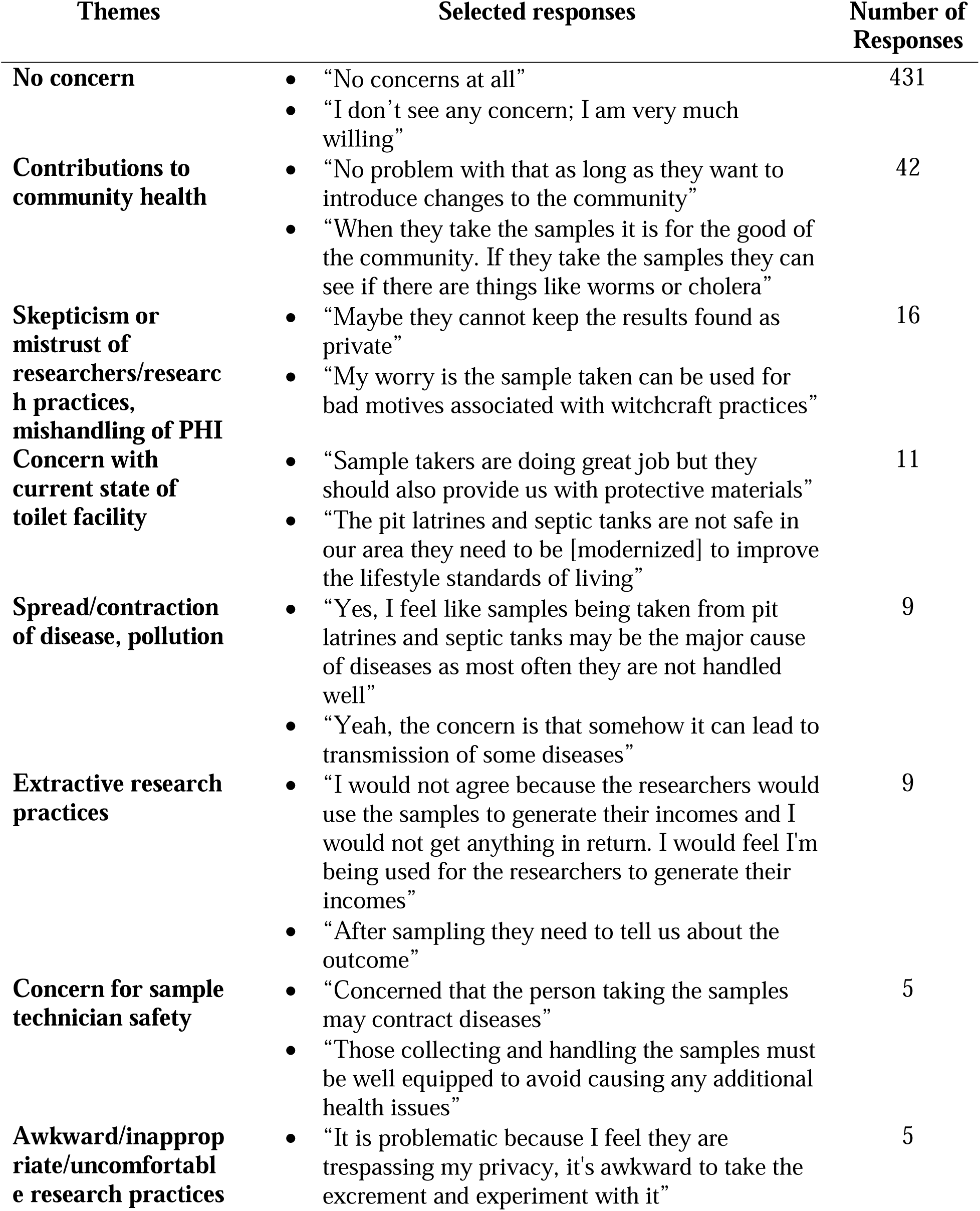
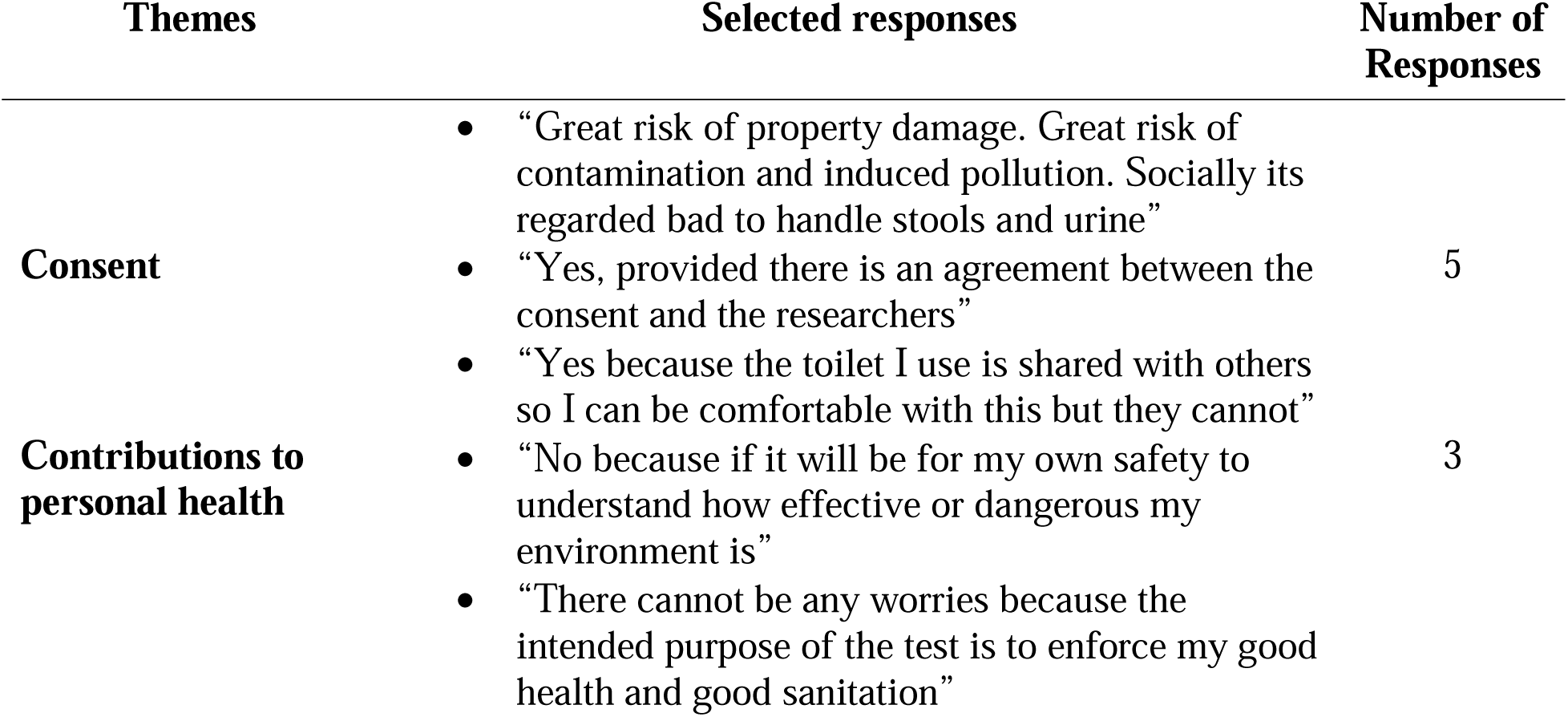
Qualitative themes and selected comments or concerns about wastewater and environmental samples being taken in a respondent’s area.

### Board game

The full board game results are presented in Supplemental Material Table S1. Approximately half (*n =* 16) of the 30 participants who played the board game completed all seven rounds. On average, participants completed five of the seven rounds, retained 40% of their privacy, and had 71% community health and 74% personal health information at the end of the game. When asked which incentives were most influential in their decision to accept the offers in the board game and give up their privacy, 13 participants mentioned incentives that benefitted their personal wellbeing, nine mentioned incentives related to community health, and seven mentioned benefits to both their personal health and the community. For the full public opinion survey, none of the board game participants indicated any concerns about samples being taken from pit latrines and flush toilets in the area. When asked which privacy-reducing measure(s) they were most concerned about, the responses were more variable, with participants commenting on losing the ability to determine who accesses their data (*n =* 5), giving away too much information (*n =* 7), lab testing/surveys (*n* = 3), wastewater monitoring and household/city-level concerns (*n* = 4), and no concerns (*n* = 11). Fourteen board game participants decided to reject the offer presented to them before reaching round seven. When asked why they decided to do so, eight stated that they wanted to retain their privacy, while others believed that their requested information was too personal or was being shared with too many people.

In the full survey, respondents who also played the board game and responded that they viewed WES as an invasion of privacy (*n* = 7), on average, retained 41% of their privacy in the board game, compared to the 39% who responded that they did not view WES as an invasion of privacy.

## Discussion

WES researchers, and those who make public health decisions with WES data in LMIC communities and camps, have access to information about the health risks and status of the community, which may inform decisions on how best to improve the health of the individuals where samples are being obtained. While physicians must keep this information private, WES data can be collected by the government, local consultant research groups, academic researchers, NGOs, or locally elected officials. Many survey respondents were willing to participate in WES; however, their desire to know about the results was high. This factor may be more important in Malawi than was previously thought. In terms of the African “ubuntu” concept,^18,19^ WES used in geographically grouped settings fits a communal, non-individualistic model, and this cultural cornerstone could be the root of high WES support of pooled samples and few concerns by Malawian or Malawi camp residents.

We found general support for the use of WES for public health surveillance across Malawi. The most recent results by Afrobarometer, a public opinion survey of democracy, governance, economic conditions, and related issues, reported a Malawian opinion that health was among the most important problems facing the country and that the government should address.^34^ Literature also indicates the general comfort of respondents when their participation in health research is engaged.^20,35^ Our study was conducted at locations where WES samples were actively collected. Respondents were supportive of samples being collected to improve community health, and a few participants commented on improvements to their personal health as a result of wastewater monitoring. The potential to provide future benefits to society and/or to themselves has also been found to positively influence participation,^20,36^ this could be extended to include the desire to answer important research questions that WES may address.

Although most respondents did not express concerns, some did. Themes in these concerns included pollution, the spread of diseases, the safety of the sample takers, extractive research practices, general comfort regarding sample taking, mistrust of researchers/research practices, the current state of their toilets being sampled, and consent around non-sewered sanitation systems that may be used by others. Additionally, the participants wanted to be informed of the results of wastewater sampling, which is consistent with the literature indicating that receiving a copy of the results of participation in health research positively influenced the willingness to participate.^37^

Establishing a high level of trust between the public and surveillance researchers as well as between trusted messengers and data users is essential in WES. The respondents most supported the government and academic researchers using their health research data. This trust was not extended to local officials (neighborhood block leaders). This low trust at the local versus national level is consistent with other public-health trust studies conducted in the United States.^38^

Trust has been identified as an important factor in willingness to share PHI, with participants with high levels of distrust in the healthcare system being less willing to share their information.^35^ Contrastingly, in Canada, Teschke et al.^37^ found that respondents were more willing to participate in health research conducted by universities and hospitals as opposed to private research firms, private health groups, public health departments, and other government entities. Although higher-resource countries are thought to emphasize individualism more, similar sentiments were expressed among the Malawian and camp residents; respondents most often regarded protecting their own health to be of utmost importance, and generally, their willingness to give up information to ensure that others could live safe and healthy lives did not supersede their confidence in officials keeping the same information private. The implication is that trust remained an important consideration in WES. To maintain public trust in WES, implementation should focus on ensuring partnering among government, local consultant research groups, academic researchers, and NGOs undertaking this work, with less emphasis on elected local officials. Public-health trust research needs further investigation in LMICs, especially considering the common use of traditional medicine providers.

Privacy is inherently linked to the disclosure of sensitive information, as the negative consequences stemming from it are often the main risk.^22,39^ Widely, the participants did not view sanitation system waste monitoring as an invasion of privacy, and 80% had no significant concerns about samples being taken from pit latrines or septic tanks in their area. Those who viewed WES as an invasion of privacy were slightly less willing to give up their health/medical and lifestyle/behavior information for the sake of others. Financial information was regarded as the most sensitive among participants, and although it is not a direct WES parameter surveillance, support for nutrition was the highest target, and nutrition could also be an indicator of household wealth.^40^ In a recent United States survey, wastewater nutrition surveillance received some support (37%) but was among the lowest rated categories;^14^ conversely, 85% of Malawian and Malawi camp residents in our survey endorsed this target of wastewater surveillance. Privacy retention was also the most influential factor, resulting in respondents’ decisions to end the board game before completing all the rounds. Playing the board game also raised concerns for participants about WES and privacy, although none expressed concerns in their initial survey responses. Overall, the respondents were most willing to disclose their health and medical information to others. This finding holds promise for the government’s application of WES in Malawi for public health monitoring and community-wide interventions.

Notions of privacy have cultural relevancies. There are contextual differences between high- and low-income countries in terms of sociocultural privacy and idealism views, as well as dominant household toileting systems for WES sampling sites.^18,7^ One of the clearest cultural differences in privacy from our study was that three times more respondents in our survey than in respondents from the United States^14^ supported household-level WES surveillance (89% and 19%, respectively). Findings across the three studied Malawi locations suggested the most support and least direct opposition for wastewater monitoring at schools (81%), whereas only 32% of United States^14^ respondents favored such monitoring at schools. Furthermore, alcohol consumption surveillance received lower support in the United States,^14^ despite alcohol consumption being higher in the United States than Malawi (9.57 liters of pure alcohol consumed per United States adult over a calendar year, compared to 3.24 liters in Malawi^41^). This indicated that, although some clear cultural differences in WES privacy exist, boundaries of surveillance targets may be needed. Regarding the importance of health data remaining anonymous/protected, we saw differences in responses at the camp versus in the total survey, possibly because respondents were less anonymous in day-to-day life than most, as they were forced to flee and may be an enemy of their home country.

There is a need to bridge the gap between ongoing WES research and the public opinion of such research, specifically, by obtaining further insights from LMIC implementation social science studies to ensure progress towards global health goals for expanding WES in LMICs. Studies on the legal and ethical implications of wastewater monitoring have mostly focused on high-income countries).^24^ Other national surveys have indicated that more than half of Malawians say that the current government has not successfully improved basic health services,^34^ and respondents may want the government to have more data so that they can do better. Researchers should work towards minimizing extractive research practices and emphasizing participants’ right to know about their community health status, while weighing this consideration against best practices for improving their health. It is recommended that researchers do not make assumptions regarding the WES-related opinions of individuals in LIMICs. Additional research is needed in resource-limited settings, such as those of many populations living in LMICs, to better understand public opinion and develop best practices for sharing WES data with participants.

### Limitations

The findings may not be generalizable to the overall population of Malawi or to other LMICs, because the self-reported data were collected from a convenience sample rather than a random sample. The data overrepresented men and the urban areas, particularly staff and students from higher-education campus facilities. Not all surveys could be conducted in the native language of the refugees and asylum seekers, owing to logistical challenges and constraints provided by the ethical review boards; however, many spoke English or Chichewa. Because refugees and asylum seekers have sociodemographic characteristics different from the national population, weighted proportions were not calculated in this study. Therefore, it was not possible to deduce whether the differences observed between the Malawians and Dzaleka camp residents were driven by their refugee status or by their home culture/nationality. There were other privacy factors, outside of PHI, that may have prompted individual concern such as those related to illegal drug surveillance, as well as related to policing or concerns about predatory health insurance practices in areas with high alcohol use or smoking; although these concerns have been documented in the United States, they may be less of an issue in LMICs. Additionally, in the board game, participants were presented with hypothetical scenarios. It is possible that board game actions reflected a stronger commitment to privacy than would be present in the same or similar real-world situations. When viewing the consistency of the questions, we found that the instrument had good reliability.

## Conclusion

Overall, our work suggested that the Malawian and Malawi camp residents supported ongoing WES for public health and did not have many PHI or privacy concerns. Higher support was found for examining targets such as communicable diseases, deadly diseases, environmental toxicants, healthy eating, illegal drugs, mental illnesses, and prescription drugs, and when WES was conducted in the entire city or at camps and schools. There were some concerns for lifestyle behavior and alcohol surveillance, as well as for surveillance conducted at businesses and religious organizations. Importantly, respondents consistently expressed the desire to have WES data communicated back to them; thus, developing effective WES communication approaches for LMICs is a critical gap in this field, and leaving insufficient time to gauge public perception could be a mistake for other ongoing WES efforts in LMICs. Communication efforts may include community sensitization to raise the awareness of those living where sampling is being conducted, or programs such as children’s science day camps.

Our study is the first to examine the acceptance of WES research in an LMIC, including the opinions of refugees and asylum seekers. Considering privacy thresholds and participant autonomy regarding public health and access among cultural relevancies is important for future policy development and investment. We found that the acceptance of targets and geographic scales for WES varied between this study conducted in Malawi and previous studies performed in the United States. This finding is particularly helpful for the surveillance of marginalized and politically vulnerable populations, such as refugees whose opinions in our survey differed in some cases from the overarching Malawian opinion. This study illustrates the benefits of adding public opinion inquiries as a social science extension where active WES sample collection is underway in LMICs. While such a survey should be contextualized (for example, sewered versus non-sewered sanitation system catchment area questions may differ), community responses to a core set of questions would ultimately be valuable. Such questions include: 1.) What are the community views regarding WES in this area? 2.) What targets are acceptable to monitor? 3.) What are the population catchment size boundaries? and 4.) Should the data results be communicated back?

## Data Availability

All data generated or analyzed during this study are included in this published article and its supplementary information files.

## Acknowledgements

We thank Promise Kayala, John Ngandu, and Aubrey Dzinkambani for their assistance with translation.

## Authors’ contributions

Conceptualization: DJ, RHH; Methodology: DJ, RHH; Formal analysis: DJ, RHH; Writing-original draft preparation: DJ, RHH; Writing-review and editing: DJ, BBS, PC, BAC, AE, CW, JLH, TS, RHH; Supervision: RHH. All authors have read and agreed to the published version of the manuscript.

## Supplementary Material

Supplementary Material Text S1 - Survey on public perceptions regarding acceptance of use of wastewater for community health monitoring in Malawi (English)

### Demographics

1. What gender do you identify as?

Male
Female
Non-binary / third gender
Prefer not to say
Other
2. What is your age range?

1. 18 – 24
2. 25 – 34
3. 35 – 44
4. 45 – 54
5. 55 – 64
6. 65 – 74
7. 75 – 84
8. 85 or older
3. What is the highest level of education you completed?

Primary school
Secondary school
Some University
University degree
Technical degree
4. What type of toilet do you use most often:

Pit latrine
Flush toilet to a septic tank
No facility/Open defecation

### Block A

1. Both the Ministry of Health and Ministry of Water and Sanitation conduct environmental activities to protect the public’s well-being. If it were possible, would you be comfortable with the monitoring of sanitation system waste for the following items? <u>For each activity</u>, use the corresponding number(s) below to indicate your level of support or opposition. Very comfortable **- 5** Somewhat comfortable - **4** Neutral – **3** Somewhat uncomfortable – **2** Very uncomfortable – **1**

a. Illegal drugs
b. Prescription drugs
c. Alcohol
d. Environmental toxins (e.g., industrial chemicals)
e. Deadly diseases (e.g., Ebola, Tuberculosis)
f. Communicable diseases (e.g., Cholera)
g. Mental illness (e.g., stress hormones)
h. Lifestyle behaviors (e.g., smoking, birth control)
i. Healthy eating

### Block B

1. If this area were to monitor sanitation systems, what would you want monitored? Check all that apply

a. The entire city/camp
b. Neighborhoods
c. Businesses
d. Prisons
e. Schools
f. Houses
g. I would not support any monitoring of sanitation system waste
2. Which of the following areas, if any, should be prohibited from sanitation system waste monitoring? Check all that apply.

a. Religious organizations
b. Schools, Colleges and Universities
c. Individual households
d. Private companies
e. I would support monitoring of all of these places
3. Would you view conducting sanitation system waste monitoring as an invasion of privacy?

a. No, it is not an invasion of privacy
b. Yes, it is an invasion of privacy
c. I don’t know

### Block C

1. How much would you support the following groups accessing your health data? Use the corresponding number(s) below to indicate your level of support or opposition

Full support - **5**
Support somewhat - **4**
Neutral - **3**
Oppose somewhat - **2**
Full opposition - **1**
a) Government officials
b) Consultant research groups
c) Academic researchers
d) NGO supported researchers
e) Block leader initiatives

### Block D

Number the options 1 through 5 to show what is most important to you. 1 will be ‘most important’ and 5 will be ‘least important’.

a. Protect the health of my loved ones
b. Protect the health of people I know
c. Protect the health of people I do not know
d. Protect the health of all people in my city/camp
e. Protect my health

### Block E

If sanitation system waste was to be monitored, indicate the importance of the following items to you

Very important - **5**
Somewhat important - **4**
Neutral **- 3**
Somewhat unimportant **- 2**
Unconcerned - **1**

a. How important is it that analysis of your health data is communicated back to you?
b. How important is it to you that your health data remains anonymous/protected?

### Block F

1. How much confidence do you have in Officials to keep your personal information private? Use a number show your level of confidence (0 = No confidence, 4 = Complete confidence, or 1,2,3 can indicate somewhere in-between)

a. Health or Medical information
b. Financial information
c. Lifestyle / behaviors information
2. How much of your privacy would you be willing to give up to ensure that people in your area can live safe and healthy lives? Use a number show your level of willingness (0 = None at all, 4 = All of it, or 1,2,3 can indicate somewhere in-between)

a. Health or Medical information
b. Financial information
c. Lifestyle / behaviors information

### Block G

Do you have any concerns about samples being taken from the pit latrines and septic tanks in this area?

**Supplementary Material Table S1.**
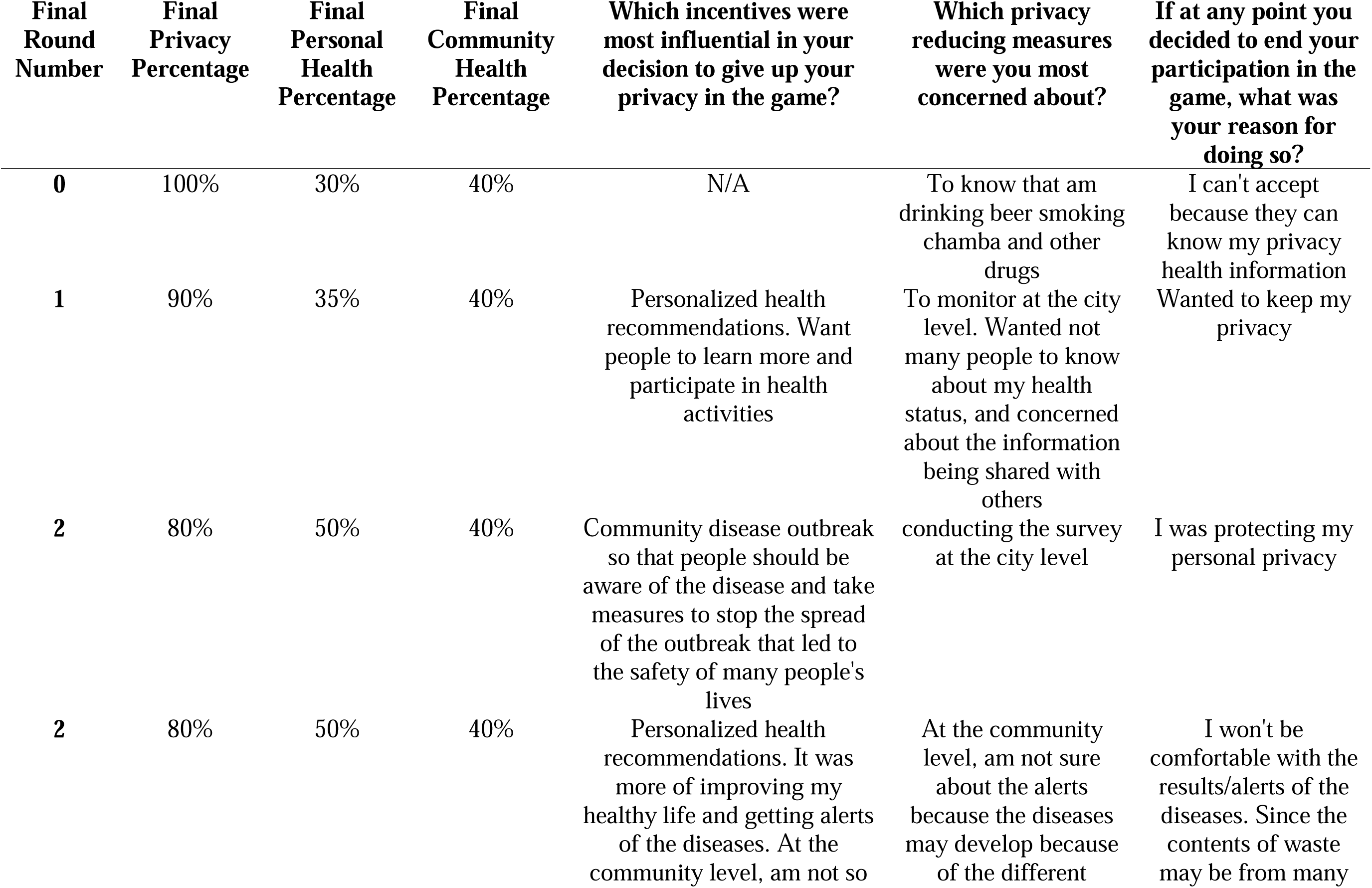
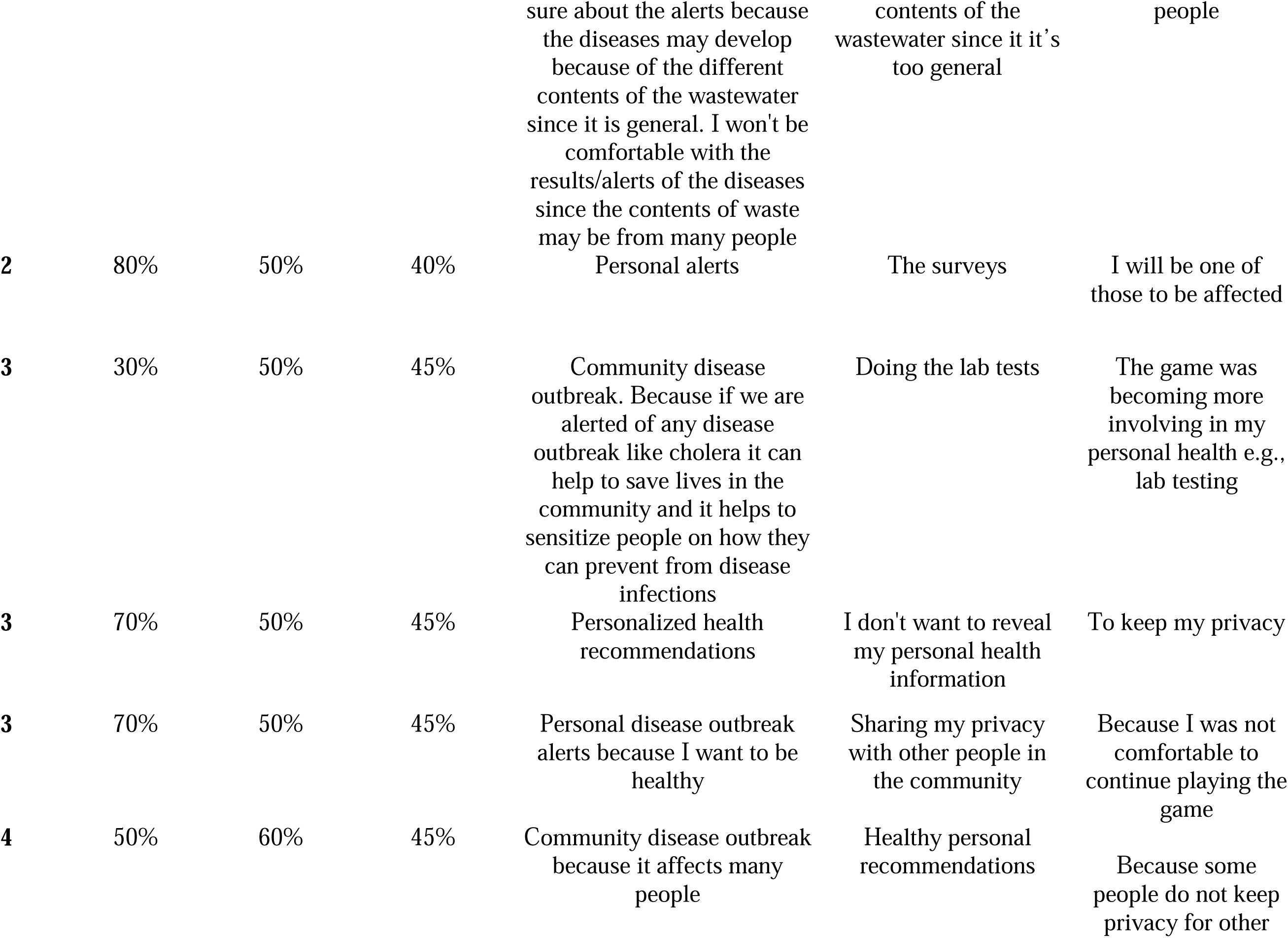
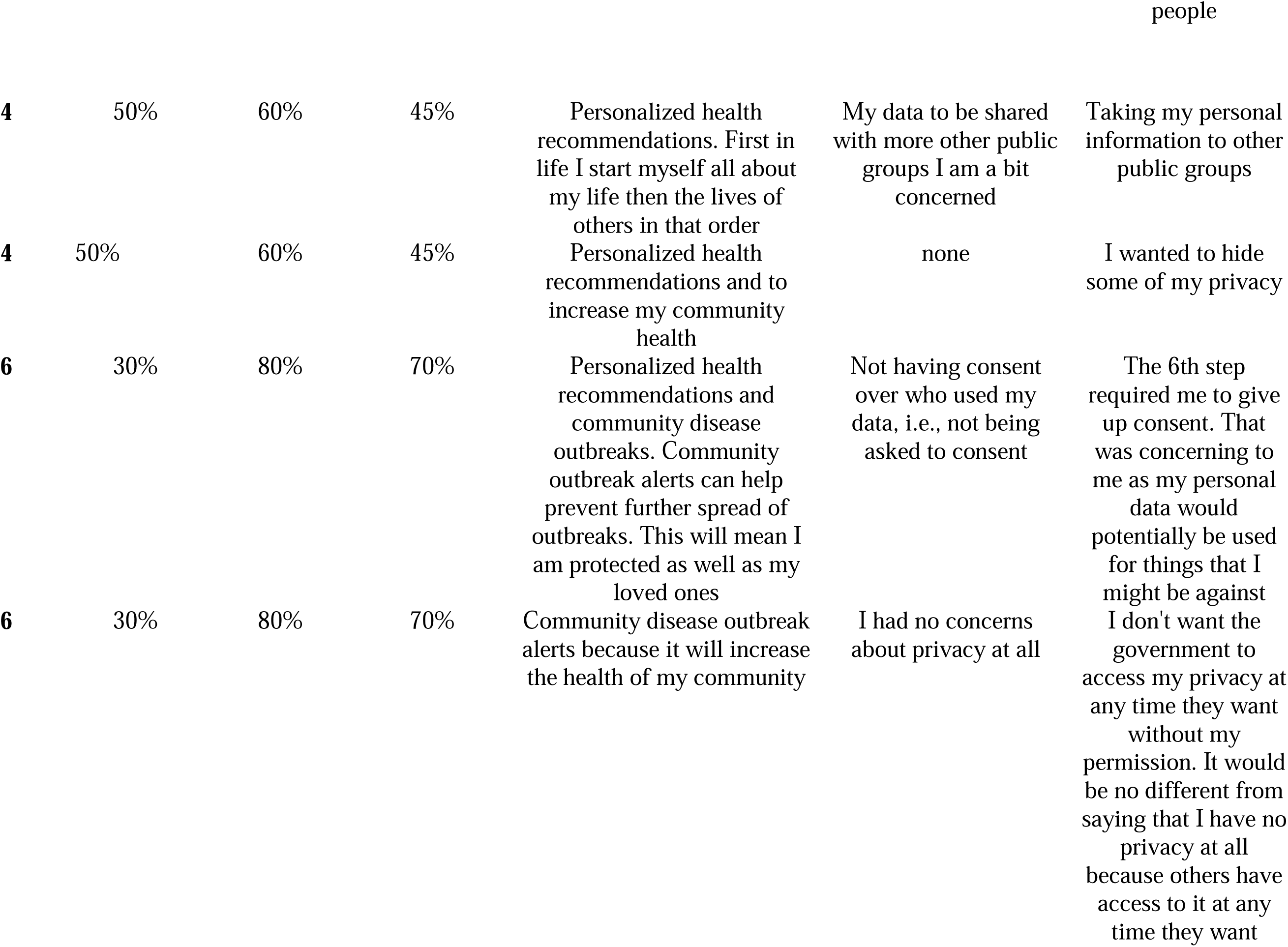
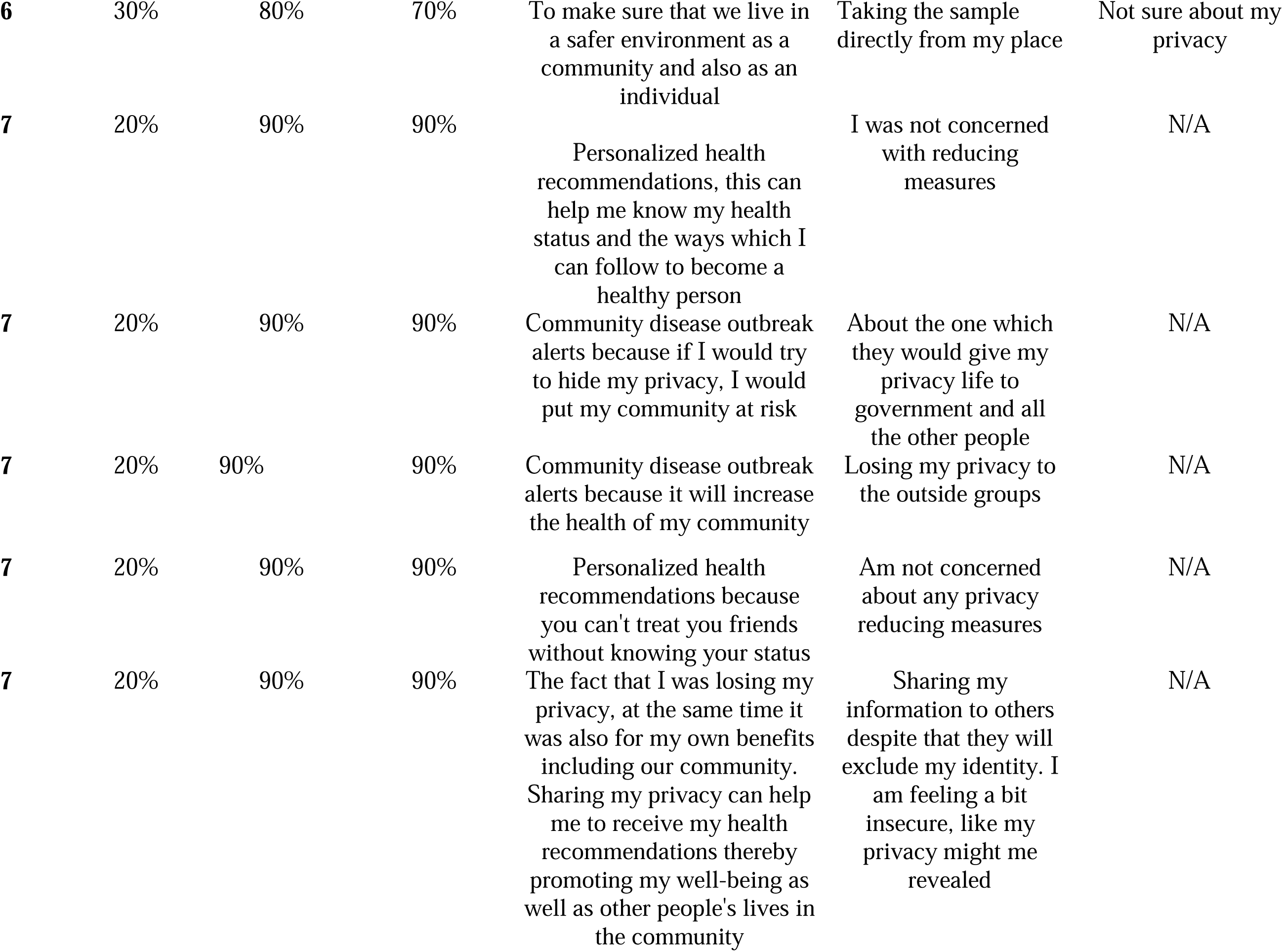
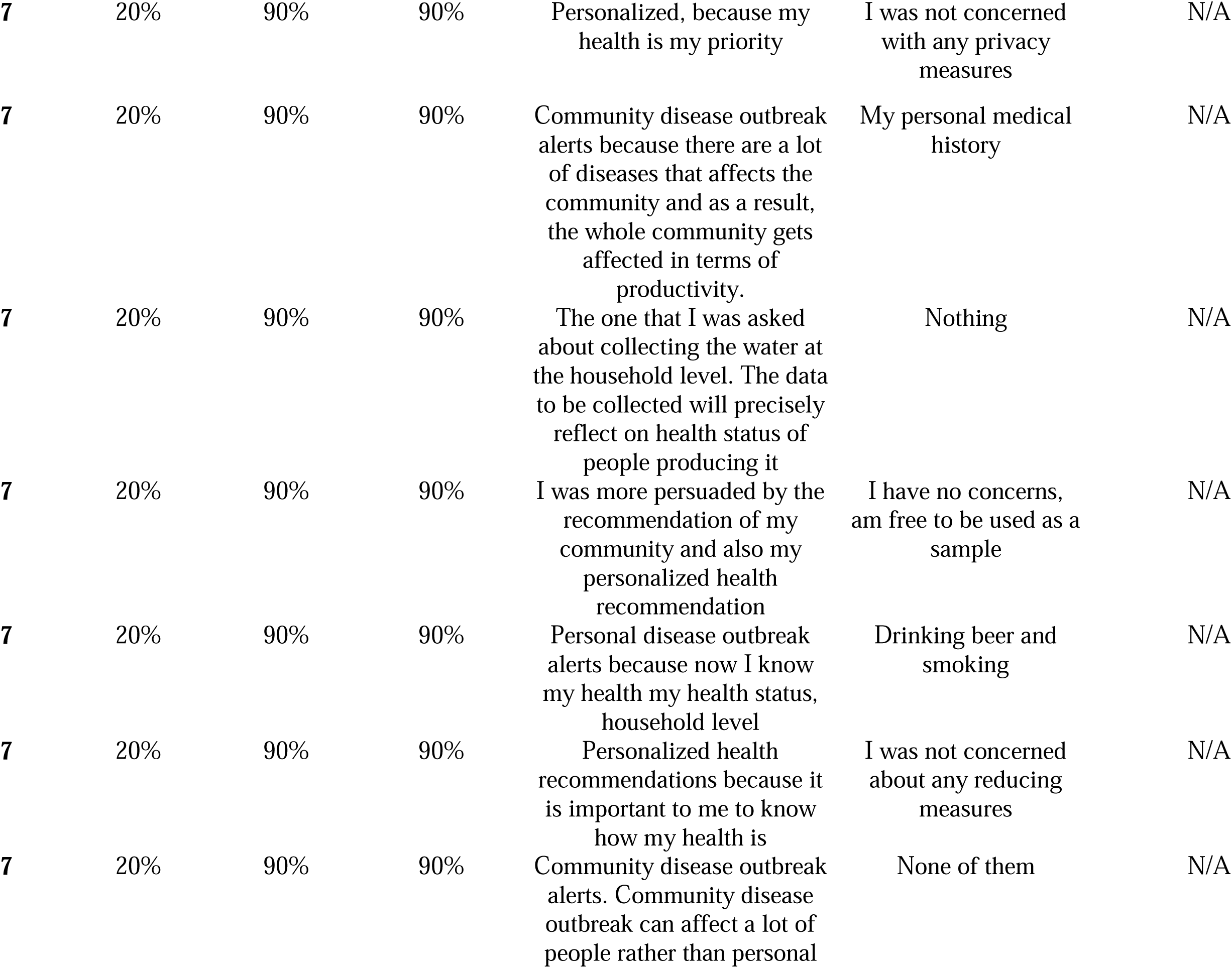
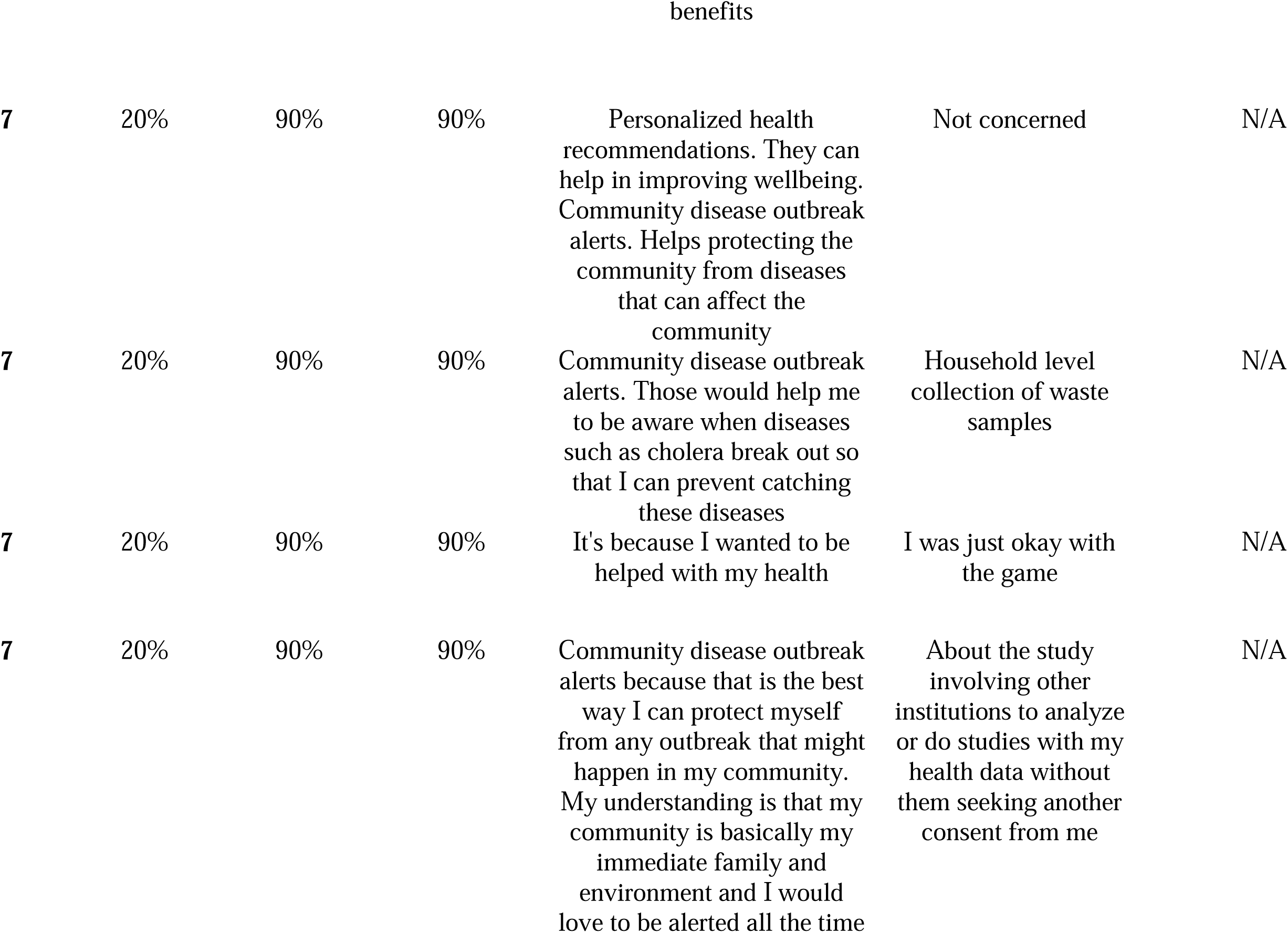
- Board game results.

| **Final Round Number** | **Final Privacy Percentage** | **Final Personal Health Percentage** | **Final Community Health Percentage** | **Which incentives were most influential in your decision to give up your privacy in the game?** | **Which privacy reducing measures were you most concerned about?** | **If at any point you decided to end your participation in the game, what was your reason for doing so?** |
| --- | --- | --- | --- | --- | --- | --- |
| **0** | 100% | 30% | 40% | N/A | To know that am drinking beer smoking chamba and other drugs | I can't accept because they can know my privacy health information |
| **1** | 90% | 35% | 40% | Personalized health recommendations. Want people to learn more and participate in health activities | To monitor at the city level. Wanted not many people to know about my health status, and concerned about the information being shared with others | Wanted to keep my privacy |
| **2** | 80% | 50% | 40% | Community disease outbreak so that people should be aware of the disease and take measures to stop the spread of the outbreak that led to the safety of many people's lives | conducting the survey at the city level | I was protecting my personal privacy |
| **2** | 80% | 50% | 40% | Personalized health recommendations. It was more of improving my healthy life and getting alerts of the diseases. At the community level, am not so | At the community level, am not sure about the alerts because the diseases may develop because of the different | I won't be comfortable with the results/alerts of the diseases. Since the contents of waste may be from many |
|  |  |  |  | sure about the alerts because the diseases may develop because of the different contents of the wastewater since it is general. I won't be comfortable with the results/alerts of the diseases since the contents of waste may be from many people | contents of the wastewater since it's too general | people |
| **2** | 80% | 50% | 40% | Personal alerts | The surveys | I will be one of those to be affected |
| **3** | 30% | 50% | 45% | Community disease outbreak. Because if we are alerted of any disease outbreak like cholera it can help to save lives in the community and it helps to sensitize people on how they can prevent from disease infections | Doing the lab tests | The game was becoming more involving in my personal health e.g., lab testing |
| **3** | 70% | 50% | 45% | Personalized health recommendations | I don't want to reveal my personal health information | To keep my privacy |
| **3** | 70% | 50% | 45% | Personal disease outbreak alerts because I want to be healthy | Sharing my privacy with other people in the community | Because I was not comfortable to continue playing the game |
| **4** | 50% | 60% | 45% | Community disease outbreak because it affects many people | Healthy personal recommendations | Because some people do not keep privacy for other |
| **4** | 50% | 60% | 45% | Personalized health recommendations. First in life I start myself all about my life then the lives of others in that order | My data to be shared with more other public groups I am a bit concerned | Taking my personal information to other public groups |
| **4** | 50% | 60% | 45% | Personalized health recommendations and to increase my community health | none | I wanted to hide some of my privacy |
| **6** | 30% | 80% | 70% | Personalized health recommendations and community disease outbreaks. Community outbreak alerts can help prevent further spread of outbreaks. This will mean I am protected as well as my loved ones | Not having consent over who used my data, i.e., not being asked to consent | The 6th step required me to give up consent. That was concerning to me as my personal data would potentially be used for things that I might be against |
| **6** | 30% | 80% | 70% | Community disease outbreak alerts because it will increase the health of my community | I had no concerns about privacy at all | I don't want the government to access my privacy at any time they want without my permission. It would be no different from saying that I have no privacy at all because others have access to it at any time they want |
| 6 | 30% | 80% | 70% | To make sure that we live in a safer environment as a community and also as an individual | Taking the sample directly from my place | Not sure about my privacy |
| 7 | 20% | 90% | 90% | Personalized health recommendations, this can help me know my health status and the ways which I can follow to become a healthy person | I was not concerned with reducing measures | N/A |
| 7 | 20% | 90% | 90% | Community disease outbreak alerts because if I would try to hide my privacy, I would put my community at risk | About the one which they would give my privacy life to government and all the other people | N/A |
| 7 | 20% | 90% | 90% | Community disease outbreak alerts because it will increase the health of my community | Losing my privacy to the outside groups | N/A |
| 7 | 20% | 90% | 90% | Personalized health recommendations because you can't treat you friends without knowing your status | Am not concerned about any privacy reducing measures | N/A |
| 7 | 20% | 90% | 90% | The fact that I was losing my privacy, at the same time it was also for my own benefits including our community. Sharing my privacy can help me to receive my health recommendations thereby promoting my well-being as well as other people's lives in the community | Sharing my information to others despite that they will exclude my identity. I am feeling a bit insecure, like my privacy might me revealed | N/A |
| 7 | 20% | 90% | 90% | Personalized, because my health is my priority | I was not concerned with any privacy measures | N/A |
| 7 | 20% | 90% | 90% | Community disease outbreak alerts because there are a lot of diseases that affects the community and as a result, the whole community gets affected in terms of productivity. | My personal medical history | N/A |
| 7 | 20% | 90% | 90% | The one that I was asked about collecting the water at the household level. The data to be collected will precisely reflect on health status of people producing it | Nothing | N/A |
| 7 | 20% | 90% | 90% | I was more persuaded by the recommendation of my community and also my personalized health recommendation | I have no concerns, am free to be used as a sample | N/A |
| 7 | 20% | 90% | 90% | Personal disease outbreak alerts because now I know my health my health status, household level | Drinking beer and smoking | N/A |
| 7 | 20% | 90% | 90% | Personalized health recommendations because it is important to me to know how my health is | I was not concerned about any reducing measures | N/A |
| 7 | 20% | 90% | 90% | Community disease outbreak alerts. Community disease outbreak can affect a lot of people rather than personal | None of them | N/A |
| benefits |  |  |  |  |  |  |
| 7 | 20% | 90% | 90% | Personalized health recommendations. They can help in improving wellbeing. Community disease outbreak alerts. Helps protecting the community from diseases that can affect the community | Not concerned | N/A |
| 7 | 20% | 90% | 90% | Community disease outbreak alerts. Those would help me to be aware when diseases such as cholera break out so that I can prevent catching these diseases | Household level collection of waste samples | N/A |
| 7 | 20% | 90% | 90% | It's because I wanted to be helped with my health | I was just okay with the game | N/A |
| 7 | 20% | 90% | 90% | Community disease outbreak alerts because that is the best way I can protect myself from any outbreak that might happen in my community. My understanding is that my community is basically my immediate family and environment and I would love to be alerted all the time | About the study involving other institutions to analyze or do studies with my health data without them seeking another consent from me | N/A |

